# Detection of SARS-CoV-2 Omicron, Delta, Alpha and Gamma variants using a rapid antigen test

**DOI:** 10.1101/2022.01.27.22269299

**Authors:** Nol Salcedo, Nidhi Nandu, Julie Boucau, Bobby Brooke Herrera

## Abstract

Throughout the coronavirus disease 2019 (COVID-19) pandemic, severe acute respiratory syndrome coronavirus 2 (SARS-CoV-2) variants have emerged with different infection and disease dynamics. Testing strategies, including clinical diagnosis, surveillance, and screening, have been deployed to help limit the spread of SARS-CoV-2 variants. Rapid antigen tests, in particular, have been approved for self-testing in many countries and governments are supporting their manufacturing and distribution. However, studies demonstrating the accuracy of rapid antigen tests in detecting SARS-CoV-2 variants, especially the new Omicron variant, are limited. We determined the analytical sensitivity of a CE-marked rapid antigen test against the Omicron, Delta, Alpha and Gamma variants. The rapid antigen test had the most sensitive limit of detection (10 plaque forming units [PFU]/mL) when tested with the Alpha and Gamma variants, followed by the Omicron (100 PFU/mL) and Delta (1,000 PFU/mL) variants. Given the increasing numbers of breakthrough infections and the need to surveil infectiousness, rapid antigen tests are effective public health tools to detect SARS-CoV-2 variants.

## Introduction

Coronaviruses (CoVs) are a diverse family of positive-sense single-stranded RNA viruses, which can infect humans and other mammals. In the past 20 years, CoVs have emerged in human populations.^1^ Human CoVs (i.e., HCoV-229E, -OC43, -NL63, and -HKU1) have long been known to circulate seasonally usually causing mild respiratory tract infections.^2–5^ In contrast, severe acute respiratory syndrome coronavirus (SARS-CoV), Middle East respiratory syndrome coronavirus (MERS-CoV), and SARS-CoV-2 are highly pathogenic.^6^

In January 2020, SARS-CoV-2 was identified as the causative agent of the coronavirus disease 2019 (COVID-19) outbreak first detected in Wuhan, China (Fig. 1).^7,8^ The World Health Organization declared COVID-19 a pandemic in March 2020, and as of January 2022, more than 300 million confirmed cases and 5.4 million deaths have been reported globally. Large-scale whole-genome sequencing of the virus has identified sequence changes, particularly in the spike protein, and the emergence of novel variants.

**Figure 1.**
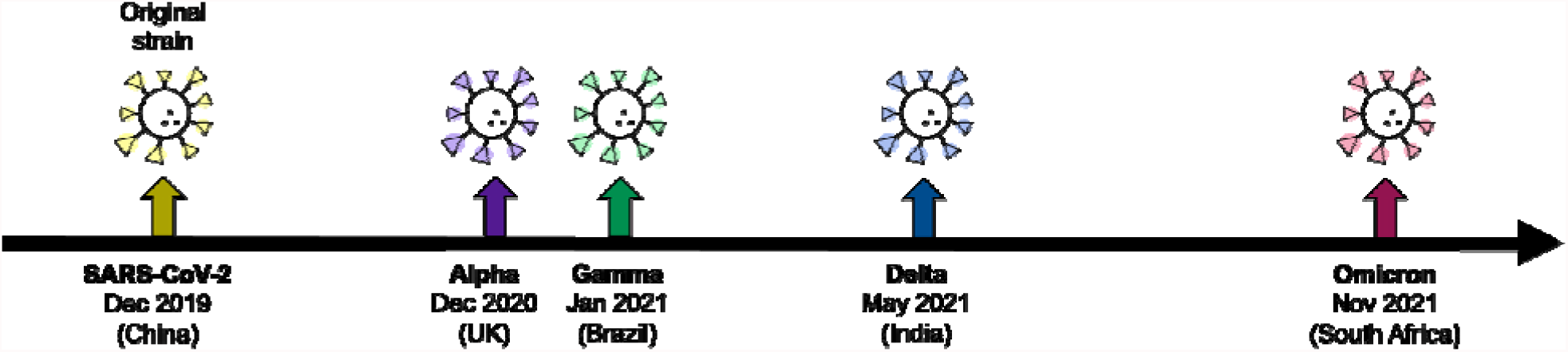
Timeline of different SARS-CoV-2 variants since the discovery of the virus. Most prevalent (<85%) nucleocapsid mutations in the variant sequences submitted to GISAID indicate mutations R203K and G204R (highlighted in red) are not common in the Delta variant.

In September 2020, the first case of infection with B.1.1.7 (Alpha) variant was identified in the United Kingdom. Genetic alterations in the Alpha variant were shown to be associated with increased binding affinity with the host cell receptor and immune evasion.^9^ Almost simultaneously, infections with B.1.351 (Beta) and P.1 (Gamma) variants were identified in South Africa and Brazil, respectively.^10,11^ In October 2020, the B.1.617.2 (Delta) variant was identified throughout India, outcompeting pre-existing variants, and establishing itself as the dominant variant until the end of 2021.^12^ Several studies have demonstrated increased transmissibility and immune evasion by the Delta variant.^12^ Additionally, COVID-19 patients infected with the Delta variant were shown to have higher risk of hospitalization, intensive care unit admission, and mortality. In November 2021, genomic surveillance in South Africa and Botswana identified infections with B.1.1.529 (Omicron) variant, and in under two months infections by the variant have been identified in 87 countries.^13^ The Omicron variant has over 30 mutations in the spike protein, influencing antibody neutralization by vaccination.

With the unprecedented spread of the Omicron variant, governments are deploying rapid antigen tests as a strategy to suppress virus transmission. However, there are limited studies that have evaluated the accuracy of antigen tests in detecting SARS-CoV-2 variants, especially Omicron. In this study, we determined the analytical sensitivity of a CE-marked rapid antigen with the Omicron, Delta, Alpha and Gamma variants. Our data indicate that despite slight differences in sensitivity, the antigen test is effective at detecting SARS-CoV-2 variants, including the dominant Omicron variant.

## Methods

### Viral isolates

The Omicron, Delta, Alpha and Gamma variant isolates were obtained from the MassCPR variant repository. In brief, the variants were isolated at the Ragon BSL3 by collection of the culture supernatant of Vero-E6 cells at 4-6 days post-infection with primary clinical specimens. The viral titer (plaque forming unit (PFU)/mL) of each viral stock was calculated by standard plaque assay using 5-fold serial dilution of the stock in on Vero-E6 cells. Genomic sequences of each variant stock were confirmed by whole genome sequencing. The collection of isolates was approved by the Massachusetts General Hospital Institutional Review Board (approval number 2019P003305).

### Sample preparation

Stocks of the clinical isolates of Omicron, Delta, Alpha and Gamma SARS-CoV-2 virus were diluted to 100,000 PFU/mL in the kit buffer (E25Bio, Inc., Cambridge, MA and Perkin Elmer, Waltham, MA). A series of 10X dilutions (100,000 to 1 PFU/mL) in the kit buffer were made for further analysis. All laboratory procedures involving the handling of the samples were carried out in a Biosafety Level 3 (BSL-3) laboratory (Ragon Institute of MGH, Harvard, and MIT).

### SARS-CoV-2 Antigen Test

The rapid antigen test (E25Bio, Inc., Cambridge, MA and Perkin Elmer, Waltham, MA) used for the study targets the SARS-CoV-2 nucleocapsid (N); the test is CE-marked. The test and the control line have immobilized antibodies that produce visible results upon interaction with antigen and the nanoparticle conjugate. 100µL of the 10X serial dilutions (100,000 to 1 PFU/mL) were applied to the antigen test in triplicates. After 15 minutes, results were scored as positive or negative and images of the tests were captured using an iPad (Apple, Inc, Cupertino, CA).

### Image Analysis

The images of the rapid antigen tests were analyzed using Image J (NIH, Bethesda, MD) for quantitative analysis of the results. The software was used to calculate the average pixels of the test line, control line and the background area. The signal from the background was subtracted from the test and control line signals before normalizing the test signal to the control signal. The resulting test signal expressed as percent of control was used to determine the limit of detection for the rapid antigen test.

### Statistics

GraphPad Prism 9.0 (San Diego, CA) was used to analyze and report the final test signal of the antigen tests. The mean value of the test signal with standard deviations were plotted in column graphs.

## Results

To determine the analytical sensitivity of a rapid antigen test with SARS-CoV-2 variants, we utilized clinical isolates of Omicron, Delta, Alpha and Gamma. Each of the variants were diluted to 100,000 PFU/mL test samples, followed by 10X serial dilutions to obtain 10,000, 1,000, 100, 10, and 1 PFU/mL test samples. The limits of detection were determined by applying to each rapid antigen test 100 μL of the test samples. The antigen tests reacted for 15 minutes before results were visually scored and images were captured.

The rapid antigen test detected the Delta variant with the highest limit of detection at 1,000 PFU/mL, followed by the Omicron variant at 100 PFU/mL (Fig. 2A-D, Table 1). The rapid antigen test had the lowest limits of detection against the Alpha and Gamma variants at 10 PFU/mL (Fig. 2A-E, Table 1). The rapid antigen test was negative when tested with the 1 PFU/mL test samples from the Alpha and Gamma variants and the kit buffer alone (Fig. 2F-G, Table 1). Image analysis of test signal intensities corroborated our visual scoring results (Fig. 3).

**Table 1.** Rapid antigen test results for the SARS-CoV-2 variants performed in triplicate.

| SARS-CoV-2 Variant | Test Samples |  |  |  |  |  |
| --- | --- | --- | --- | --- | --- | --- |
|  | 100,000 PFU/mL | 10,000 PFU/mL | 1,000 PFU/mL | 100 PFU/mL | 10 PFU/mL | 1 PFU/mL |
| Omicron | 3/3 | 3/3 | 3/3 | 3/3 | 0/3 | Not Tested |
| Delta | 3/3 | 3/3 | 3/3 | 3/3 | Not Tested | Not Tested |
| Alpha | 3/3 | 3/3 | 3/3 | 3/3 | 2/3 | 0/3 |
| Gamma | 3/3 | 3/3 | 3/3 | 3/3 | 3/3 | 0/3 |

**Figure 2.**
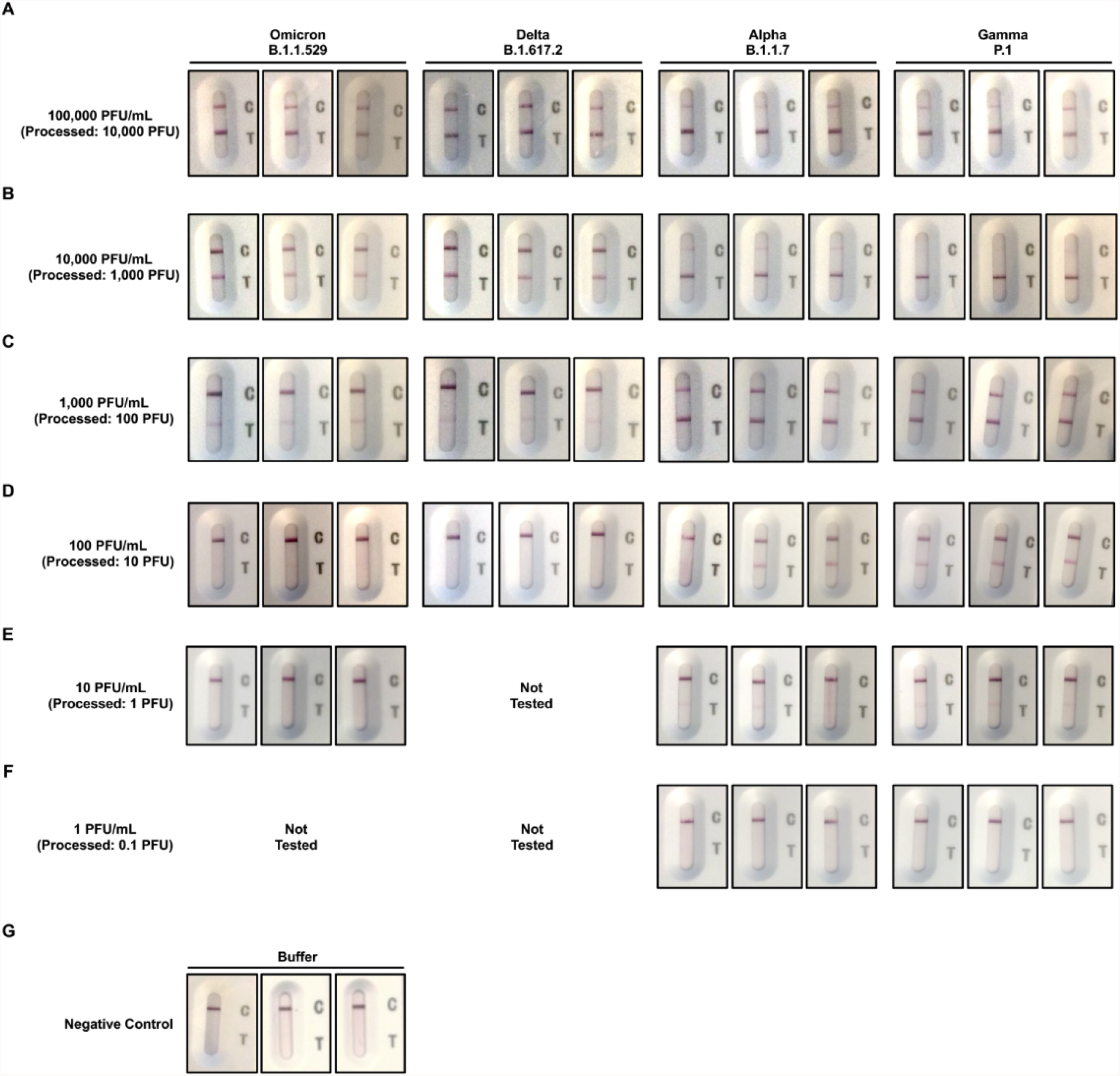
Images of rapid antigen tests for different concentrations of Omicron, Delta, Alpha, and Gamma variants recorded after 15 minutes. A) 100,000 PFU/mL. B) 10,000 PFU/mL. C) 1,000 PFU/mL. D) 100 PFU/mL. E) 10 PFU/mL. F) 1 PFU/mL. G) Kit buffer. All tests were carried out in triplicates.

**Figure 3.**
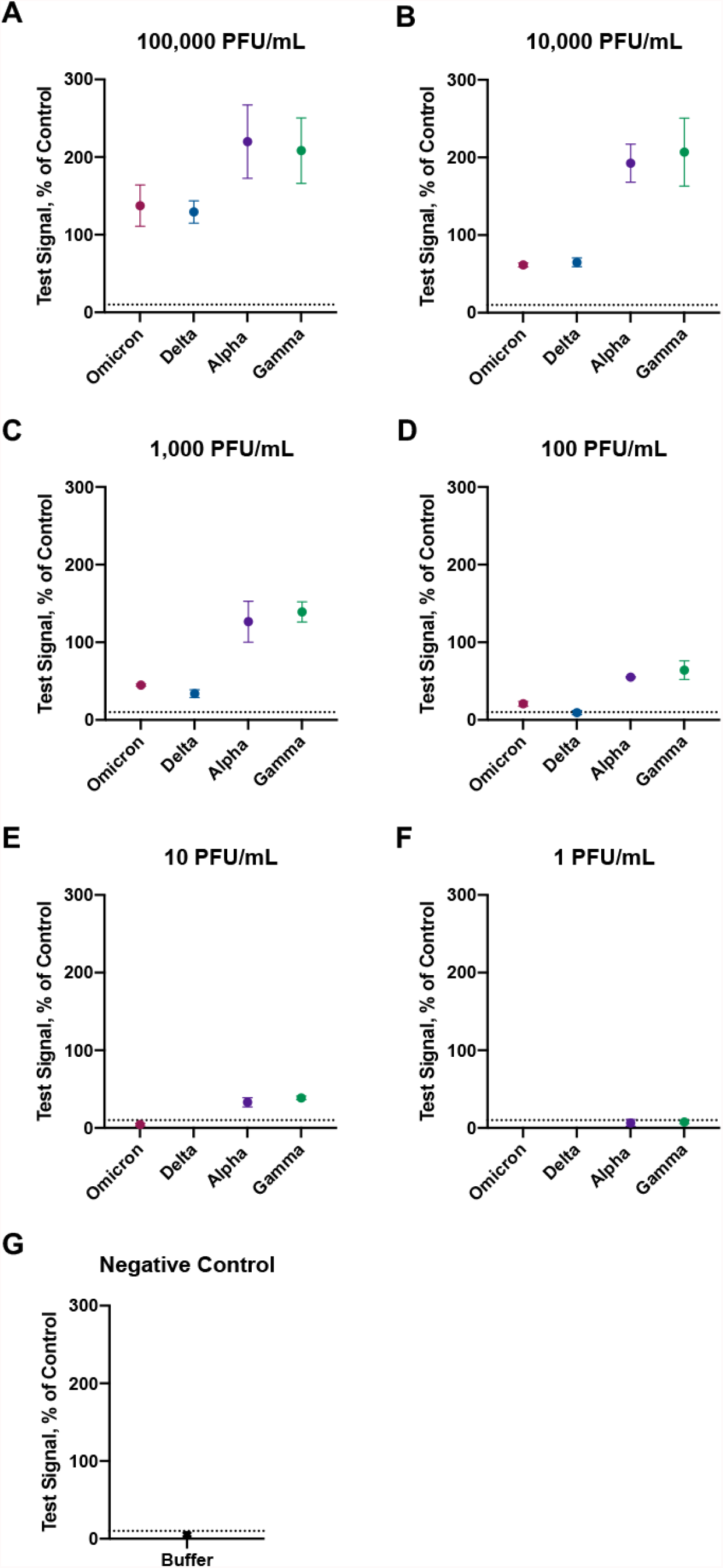
Test signals (percent of control) of rapid antigen tests for different concentrations of Omicron, Delta, Alpha, and Gamma variants. A) 100,000 PFU/mL. B) 10,000 PFU/mL. C) 1,000 PFU/mL. D) 100 PFU/mL. E) 10 PFU/mL. F) 1 PFU/mL. G) Kit buffer. The y-axis corresponds to the background subtracted test signal normalized to the control line. Test results less than 10% of the control are considered negative results, which is indicated by the black dashed line.

## Conclusions

Reverse-transcription polymerase chain reaction (RT-PCR) is the gold standard method for clinical diagnosis of COVID-19. However, due to its high cost and slow turnaround times, RT-PCR remains impractical for surveilling and screening of infectious individuals. In contrast, antigen tests can be inexpensive, self-administered, and provide rapid results. Studies have demonstrated that the sensitivity of antigen tests increases with testing frequency.^14–16^ Other studies have shown that antigen tests are optimized at detecting SARS-CoV-2 during the peak period of transmission.^15,17^ Altogether these data suggest that rapid antigen tests are optimal public health tools for surveilling infectiousness and can be used to help reduce the spread of COVID-19.

With the emergence and unprecedented expansion of the Omicron variant, governments are deploying rapid antigen tests nationally to facilitate outbreak suppression. However, there are limited studies that have determined whether antigen tests can detect the Omicron variant. Using an FDA Emergency Use Authorized antigen test, a recent study showed detection of Omicron and Delta specimens with concentrations of 100,000 copies per swab or greater.^18^ In this study, we determined the analytical sensitivity of a rapid antigen test with SARS-CoV-2 variants, including Omicron, Delta, Alpha, and Gamma. Our data show that the rapid antigen test has the lowest limit of detection with Alpha and Gamma (10 PFU/mL), followed by Omicron (100 PFU/mL) and Delta (1,000 PFU/mL) (Fig. 2-3, Table 1).

SARS-CoV-2 N is one of the predominantly expressed structural proteins and therefore is an ideal target for detection. Most of the antigen tests developed target SARS-CoV-2 N and mutations in this protein can impact the detection of the virus. In more than 85% of Omicron, Alpha, and Gamma sequences, R230K and G204R mutations in N are observed.^19^ However, those mutations were observed in less an 0.1% of Delta sequences.^19^ These data could help explain the lower analytical sensitivity of the rapid antigen test with Delta as compared to the other variants. Another possibility is that the viral stocks used in the study may contain non-infectious particles and therefore varying concentrations of N that could impact the signal intensity of the test results. Further studies are needed to understand which mutations in the N of the SARS-CoV-2 variants negatively impact the limits of detection in rapid antigen tests. Additionally, studies that evaluate the performance of the rapid antigen test using nasal or oropharyngeal swab specimens from RT-PCR confirmed COVID-19 patients are warranted. Despite slight differences in analytical sensitivity, the antigen test used in this study is effective at detecting SARS-CoV-2 variants.

## Data Availability

All data produced in the present study are available upon reasonable request to the authors

## Author Contributions

Conceptualization: BBH. Formal analysis: NS, NN, JB, BBH. Investigation: NS, NN, JB, BBH. Methodology: NS, NN, JB, BBH. Project Administration: BBH. Resources: BBH. Supervision: BBH. Validation: NS, NN, JB, BBH. Visualization: NS, NN, JB, BBH. Writing – original draft: NN, BBH. Writing – review and editing: NS, NN, JB, BBH. All authors contributed to the article and approved the submitted version.

## Funding

The authors declare that this study received funding from E25Bio, Inc. The funder was not involved in the study design, collection, analysis, interpretation of data, the writing of this article or the decision to submit it for publication.

The MassCPR variants repository is funded by the Massachusetts Consortium on Pathogen Readiness. The Ragon BSL3 is partially supported by the Harvard Center for AIDS Research, a National Institutes of Health funded program [P30AI060354] and the Massachusetts Consortium on Pathogen Readiness.

## Data Availability

All data produced in the present study are available upon reasonable request to the authors

## Conflict of Interest

NS, NN, and BBH are employed by E25Bio, Inc., a biotechnology company that develops diagnostic assays for infectious diseases.

## Notes

### Author Declarations

The collection of isolates was approved by the Massachusetts General Hospital Institutional Review Board (approval number 2019P003305).

## References

1. Ye, Z.-W. et al. Zoonotic origins of human coronaviruses. Int. J. Biol. Sci. 16, 1686–1697 (2020).

2. J., G. J. & C., W. P. Remembering seasonal coronaviruses. Science (80-.). 370, 1272–1273 (2020).

3. Abdul-Rasool, S. & Fielding, B. C. Understanding Human Coronavirus HCoV-NL63. Open Virol. J. 4, 76–84 (2010).

4. van der Hoek, L. et al. Identification of a new human coronavirus. Nat. Med. 10, 368–373 (2004).

5. Liu, D. X., Liang, J. Q. & Fung, T. S. Human Coronavirus-229E, -OC43, -NL63, and -HKU1 (Coronaviridae). Encycl. Virol. 428–440 (2021) doi:10.1016/B978-0-12-809633-8.21501-X.

6. Guarner, J. Three Emerging Coronaviruses in Two Decades: The Story of SARS, MERS, and Now COVID-19. Am. J. Clin. Pathol. 153, 420–421 (2020).

7. Zhou, P. et al. A pneumonia outbreak associated with a new coronavirus of probable bat origin. Nature 579, 270–273 (2020).

8. World Health Organization. Tracking SARS-CoV-2 variants. https://www.who.int/en/activities/tracking-SARS-CoV-2-variants/.

9. Walker, A. S. et al. Tracking the Emergence of SARS-CoV-2 Alpha Variant in the United Kingdom. N. Engl. J. Med. 385, 2582–2585 (2021).

10. Slavov, S. N. et al. Genomic monitoring unveil the early detection of the SARS-CoV-2 B.1.351 (beta) variant (20H/501Y.V2) in Brazil. J. Med. Virol. 93, 6782– 6787 (2021).

11. Gräf, T. et al. Identification of a novel SARS-CoV-2 P.1 sub-lineage in Brazil provides new insights about the mechanisms of emergence of variants of concern. Virus Evol. 7, veab091 (2021).

12. Mlcochova, P. et al. SARS-CoV-2 B.1.617.2 Delta variant replication and immune evasion. Nature 599, 114–119 (2021).

13. Raquel Viana, Sikhulile Moyo, Daniel G. Amoako, Houriiyah Tegally, Cathrine Scheepers, Christian L. Althaus, Ugochukwu J. Anyaneji, Phillip A. Bester, Maciej F. Boni, Mohammed Chand, Wonderful T. Choga, Rachel Colquhoun, Michaela Davids, Koen, A. von G. & T. de O. Rapid epidemic expansion of the SARS-CoV-2 Omicron variant in southern Africa. Nature (2022) doi:doi: https://doi.org/10.1038/d41586-021-03832-5.

14. Larremore, D. B. et al. Test sensitivity is secondary to frequency and turnaround time for COVID-19 screening. Sci. Adv. 7, (2021).

15. Harmon, A. et al. Validation of an At-Home Direct Antigen Rapid Test for COVID-19. JAMA Netw. open 4, e2126931 (2021).

16. Beatrice Nash, Anthony Badea, Ankita Reddy, Miguel Bosch, Nol Salcedo, Adam R. Gomez, Alice Versiani, G. C. D. S. and T.M.I.L. dos S. Validating and Modeling the Impact of High-Frequency Rapid Antigen Screening on COVID-19 Spread and Outcomes. J Clin Trials (2021).

17. Salcedo, N., Harmon, A. & Herrera, B. B. Pooling of Samples for SARS-CoV-2 Detection Using a Rapid Antigen Test. Front. Trop. Dis. 2, 707865 (2021).

18. Regan, J. et al. Detection of the omicron variant virus with the Abbott BinaxNow SARS-CoV-2 Rapid Antigen Assay. medRxiv (2021) doi:10.1101/2021.12.22.21268219.

19. Alaa Abdel Latif, Julia L. Mullen, Manar Alkuzweny, Ginger Tsueng, Marco Cano, Emily Haag, Jerry Zhou, Mark Zeller, Emory Hufbauer, Nate Matteson, Chunlei Wu, Kristian G. Andersen, Andrew I. Su, Karthik Gangavarapu, Laura D. Hughes, and the C. for V. S. B. Lineage Comparison.

